# Whole-exome sequencing study of opioid dependence offers novel insights into the contributions of exome variants

**DOI:** 10.1101/2024.09.15.24313713

**Authors:** Lu Wang, Yaira Z. Nuñez, Henry R. Kranzler, Hang Zhou, Joel Gelernter

**Affiliations:** Department of Psychiatry, Yale University School of Medicine, New Haven, CT; Veterans Affairs Connecticut Healthcare System, West Haven, CT; Department of Psychiatry, University of Pennsylvania Perelman School of Medicine, Philadelphia, PA; Crescenz Veterans Affairs Medical Center, Philadelphia, PA; Department of Biomedical Informatics and Data Science, Yale School of Medicine, New Haven, CT; Center for Brain and Mind Health, Yale School of Medicine, New Haven, CT; Department of Genetics, Yale School of Medicine, New Haven, CT; Department of Neuroscience, Yale School of Medicine, New Haven, CT

**Author notes:** **<u>Corresponding Authors:</u>** Hang Zhou, Department of Psychiatry, Yale School of Medicine; Veterans Affairs Connecticut Healthcare System Annex Center, room 1325, 200 Edison Rd, Orange, CT 06477, USA.; Joel Gelernter, Department of Psychiatry, Yale School of Medicine, Veterans Affairs Connecticut Healthcare System, 116A2, 950 Campbell Ave, West Haven, CT 06516, USA.

## Abstract

Opioid dependence (OD) is epidemic in the United States and it is associated with a variety of adverse health effects. Its estimated heritability is ∼50%, and recent genome-wide association studies have identified more than a dozen common risk variants. However, there are no published studies of rare OD risk variants. In this study, we analyzed whole-exome sequencing data from the Yale-Penn cohort, comprising 2,100 participants of European ancestry (EUR; 1,321 OD cases) and 1,790 of African ancestry (AFR; 864 cases). A novel low-frequency variant (rs746301110) in the *RUVBL2* gene was identified in EUR (*p*=6.59×10^-10^). Suggestive associations (*p*<1×10^-5^) were observed in *TMCO3* in EUR, in *NEIL2* and *CFAP44* in AFR, and in *FAM210B* in the cross-ancestry meta-analysis. Gene-based collapsing tests identified *SLC22A10*, *TMCO3*, *FAM90A1*, *DHX58*, *CHRND*, *GLDN*, *PLAT*, *H1-4*, *COL3A1*, *GPHB5* and *QPCTL* as top genes (*p*<1×10^-4^) with most associations attributable to rare variants and driven by the burden of predicted loss-of-function and missense variants. This study begins to fill the gap in our understanding of the genetic architecture of OD, providing insights into the contribution of rare coding variants and potential targets for future functional studies and drug development.

## Introduction

Opioid dependence (OD, DSM-IV^1^; similarly, opioid use disorder (OUD), DSM-5^2^) is a chronic relapsing disorder that causes clinically significant distress or impairment^3^. Symptoms of OD include an elevated desire to use opioids, tolerance, loss of control in intake, and a characteristic withdrawal syndrome that follows abrupt abstinence from opioids. In 2017, OD affected about 40.5 million people globally, with >100,000 opioid overdose deaths annually^4,5^. The highest prevalence of OD has been observed in the United States, where the use of opioids increased in recent decades^6^.

Studies of the mechanisms underlying OD include those of the neurocircuitry in the brain^3,7^ and molecular pathways involving opioid receptors^8^. Genetic epidemiological studies showed a moderate heritability of OD^9^. Thus, identifying genetic factors that contribute to OD risk could advance efforts to prevent, identify, and treat the disorder. To date, genome-wide association studies (GWAS) of OD^10-17^ have identified more than a dozen common variants significantly associated with OD in genes encoding potassium ion channels, glutamate receptors, and opioid receptors, improving our understanding of the genetic architecture of the disorder^18,19^.

The largest GWAS of OD/OUD to date was conducted in European and African ancestries subjects^17^ (EUR and AFR), identifying independent risk loci including the *OPRM1* and *FURIN* genes – both identified in other genetic studies^13,15,16^. By incorporating GWAS data of problematic alcohol use and cannabis use disorder using a multi-trait analysis of GWAS (a method that boosts power for gene discovery for a set of similar traits), 19 independent loci were identified in total, reflecting a shared genetic architecture among substance use disorders^20,21^. Another study used data from the Million Veteran Program (MVP) and identified 14 loci associated with OUD^15^. Convergent findings from these two studies include risk variants in or near genes *OPRM1*, *FURIN*, *RABEPK*, *NCAM1*, and others.

However, there remain large gaps in our knowledge of the genetics of OD. The estimated single-nucleotide polymorphism (SNP)-based heritability (*h*^2^) of OD ranges from 6.0% in the Partners Biobank^14^ to 12.8% in MVP-dominated studies^13,15-17^. When using a stringent definition of cases in MVP, requiring at least one inpatient or two outpatient ICD-9/10 OUD diagnostic codes, the *h*^2^ was 19.8% in AFR participants and 15.3% in EUR participants^15^. These estimates are all less than the estimates from genetic epidemiology, reflective of heritability that is unaccounted for in GWAS, which is usual for complex traits. Recently, whole-exome sequencing (WES) and whole-genome-sequencing data have been used to augment SNP-array data for many diseases and traits^22-24^. Both these methods outperform SNP-array-based studies in identifying rare variants and accounting for missing heritability^23,25,26^. However, there is little information in the literature concerning rare coding variants in OD^27^. Thus, a large-scale WES study is needed to provide adequate statistical power to detect rare OD variants. Here, we report findings from the largest multi-ancestry WES study in OD to date, which was conducted in the Yale-Penn cohort (YP)^10,28^ in a total of 3,890 participants (2,185 cases).

## Materials and methods

### Ethics

The Yale–Penn study was approved by the IRBs at all participating clinical sites. All participants provided written informed consent.

### Yale-Penn cohort

As described previously^28^, 4,530 samples from Yale-Penn were sequenced in four batches. Batch 1 whole-exome sequencing (WES) data were generated on the Illumina HiSeq 2000 platform with NimbleGen SeqCap exome capture V2 kit. Batch 2─4 WES data were obtained using the NovaSeq sequencing system and the xGen Exome Research Panel v1. Variants were called using the BWA-GATK pipeline and mapped to the human reference genome build 38 (hg38)^29,30^. Variants in low-complexity regions, with missingness rates>0.05, or that failed Hardy-Weinberg Equilibrium expectations (*p*<10^-6^) were filtered out.

Samples with inconsistencies in self-reported and genetic sex, heterozygosity rate outside the mean ±3 standard deviations (SD) range, total missingness rate>10%, mean sequencing depth <20, or with mean genotype quality score <55 were not retained for analysis. Furthermore, samples with a transition/transversion ratio, number of called variants, number of singletons, heterozygous/homozygous ratio, and insertion/deletion ratio outside the per-batch mean ±3SD range were removed. For the remaining samples, principal component analysis^31^ was used to assign the genetic ancestry for each sample using 1000 Genomes phase 3 data^32^ as reference, resulting in 2,102 EUR individuals and 1,790 AFR individuals.

The diagnosis of OD (DSM-IV opioid dependence) was obtained using the Semi-Structured Assessment for Drug Dependence and Alcoholism (SSADDA)^33^. Controls self-reported exposure to opioids (by answering ‘yes’ to ‘Have you ever used any of the following opiate/drugs?’). We included 1,321 EUR cases and 779 controls and 864 AFR cases and 926 controls in the downstream analyses.

### Variant annotation

Variants that passed the quality control steps were annotated using ANNOVAR^34^, which annotates functions of genetic variants utilizing update-to-date information. Variants predicted as frameshift mutations, stop gain, stop loss, or splicing site alterations were categorized as loss of function (LoF) variants. Missense variants with predicted REVEL^35^ score >0.5 were considered deleterious missense (Dmis) variants. Other missense (Omis) variants were those with a REVEL score ≤0.5.

We also annotated the functions of the top associated variants and genes using a series of bioinformatics tools, including those incorporating recent deep-learning methods. The tools include Combined Annotation-Dependent Depletion (CADD) score^36^ for variant deleteriousness (higher values indicate more deleterious cases, a commonly used cutoff for which is 20), SpliceAI^37^ to predict the splice-altering consequence (scores range from 0 to 1), and AlphaMissense^38^ to predict the pathogenicity of missense variants (scores range from 0 to 1).

### Single-variant association analysis

Single-variant association analyses were performed on variants with minor allele counts (MAC) ≥5 using SAIGE-GENE+^39^, correcting for age, sex, sequencing batch, and 10 principal components (PCs). Sex-stratified analyses in males and females were also performed, correcting for all covariates other than sex. Analyses were performed in EUR and AFR separately, followed by a cross-ancestry meta-analysis using METAL^40^ with a sample-sized weighted approach. Bonferroni correction was used to define the genome-wide significance of the associations (*p*<0.05/253,281=1.97×10^-7^).

### Gene-based tests

Rare variants may act in aggregate and therefore be detected by gene-based analysis. To identify genes associated with OD, we performed gene-based tests using SAIGE-GENE+ that incorporates multiple minor allele frequency (MAF) cutoffs and functional annotations to improve power^39^. For each gene, coding variants were aggregated by different annotations (LoF only, LoF+missense, LoF+missense+synonymous, synonymous) and by different MAF cutoffs (MAF≤0.01%, MAF≤0.1%, MAF≤1% and all). The covariates used were the same as those in single-variant analyses. EUR and AFR were analyzed separately, followed by a cross-ancestry meta-analysis^40^. The Bonferroni-corrected significance threshold for the exome-wide, gene-based analysis was set at *p*=2.90×10^-6^ (17,252 genes).

### Gene set enrichment analyses

Enrichment analyses of Gene Ontology (GO) terms^41^ and Reactome pathways^42^ were performed in g:Profiler^43^ with nominally significant (*p*<0.05) genes derived from the cross-ancestry, gene-based meta-analysis. For network visualization of the gene set enrichments, we downloaded the gene set lists of GO terms and Reactome pathways from g:Profiler and used the terms with adjusted p-value <0.01 as input for the Enrichment Map^44^ app implemented in Cytoscape^45^. Enriched terms were clustered based on the Jaccard similarity coefficient of 0.6, with singleton clusters discarded. Within-ancestry gene-set enrichment analyses were performed using selected genes (*p*<0.05) from gene-based association results of EUR and AFR, respectively. The input genes were ranked based on their respective beta values of the gene-based association tests from each ancestry. Top five terms from GO biological processes and KEGG pathways were visualized using clusterProfiler^46^ package in R.

## Results

### Single-variant association analysis

Variants were called in 4,530 WES samples using the BWA-GATK pipeline^29,30^. After quality control, there were 1,321 OD cases and 779 exposed controls in EUR (1,189,765 variants) and 864 cases and 926 exposed controls in AFR (1,189,609 variants), totaling 3,890 individuals (Table 1). In EUR, 123,324 variants with MAC ≥5 were included in single-variant association analysis using a logistic mixed model implemented in SAIGE-GENE+^39^; in AFR, 213,891 variants were included.

**Table 1.** Characteristics of the studied samples in Yale-Penn Cohort. Numbers of variants in different categories in each sample are listed. EUR, European; AFR, African; MAC, minor allele count; MAF, minor allele frequency; LoF, loss-of-function.

|  | EUR<br>(N=2,100) | AFR<br>(N=1,790) |
| --- | --- | --- |
| Age (mean±SD) | 37±11 | 43±9 |
| Female (%) | 755 (36.0) | 635 (35.5) |
| Cases | 1,321 | 864 |
| #Variants |  |  |
| Singleton | 374,384 | 319,831 |
| MAC$\geq 5$ | 121,786 | 211,176 |
| MAF$\geq 0.01$ | 57,336 | 103,067 |
| LoF | 7,921 | 7,482 |
| Missense | 259,413 | 272,956 |
| Synonymous | 159,240 | 184,327 |

In EUR, we identified a novel rare LoF variant in the *RUVBL2* gene associated with OD (rs746301110, beta=-2.60, SE=0.43, *p*=6.59×10^-10^, Supplementary Table 1). To confirm the sequencing quality of this variant, we extracted the quality metrics from the raw sequencing data. This indicated that the average depth (DP) was 56 and the average genotype quality (GQ) was 89, which both pass the variant QC thresholds (DP ≥10; GQ ≥20)^28^.

In EUR, a suggestive association (*p*<1×10^-5^) was observed for rs765832505 (*p*=8.22×10^-6^) in the *TMCO3* gene (Supplementary Table 1). In AFR, suggestive associations include rs804269 (*p*=4.15×10^-6^) in *NEIL2* and rs16860800 (*p*=5.45×10^-6^) in *CFAP44* (Supplementary Table 2). The cross-ancestry meta-analysis combining both EUR and AFR samples identified additional suggestive associations, which include two variants in the *FAM210B* gene ─ rs6099114 (*p*=5.55×10^-6^) and rs6099115 (*p*=5.75×10^-6^), which are in high linkage disequilibrium in both populations (*r*^2^ > 0.99) (Table 2, Supplementary Table 3). Sex-stratified single-variant association analyses (Supplementary Tables 4-9) identified suggestive variants, including a rare missense variant rs34270544 in females (*p*=3.54×10^-5^) located in *RHOD*, one of the Rho GTPase proteins (Supplementary Table 9). The Rho GTPase family has been reported to have potential therapeutic functions in substance use disorders^47,48^. Furthermore, rs34270544 has also been associated with obesity-related metabolic dysfunctions^49^.

**Table 2.** Top associated variants (*p*<1×10^-5^) in the single-variant association analyses. Beta and SE are presented for single-ancestry results, Zscore is presented for cross-ancestry meta-analysis results. A1, allele 1 or effective allele; A2, allele 2 or non-effective allele; Freq_A1, frequency of allele 1; EUR, European; AFR, African.

| SNP | Chr:pos (hg38) | Gene | A1 | A2 | Ancestry | Freq_A1 | Beta/Zscore | SE | P-value | Effect | CADD | SpliceAI | AlphaMissense |
| --- | --- | --- | --- | --- | --- | --- | --- | --- | --- | --- | --- | --- | --- |
| rs746301110 | 19:49010483 | *RUVBL2* | - | ATAGA | EUR | 0.0064 | -2.60 | 0.43 | $6.59 \times 10^{-10}$ | LoF | 27.0 | 1.00 | - |
| rs804269 | 8:11771610 | *NEIL2* | C | T | AFR | 0.46 | 0.32 | 0.07 | $4.15 \times 10^{-6}$ | noncoding | 0.38 | 0.06 | - |
| rs16860800 | 3:113294658 | *CFAP44* | A | G | AFR | 0.02 | -1.10 | 0.25 | $5.45 \times 10^{-6}$ | noncoding | 3.48 | 0.02 | - |
| rs6099114 | 20:56366068 | *FAM210B* | C | T | EUR+AFR | 0.66 | -4.54 | | $5.55 \times 10^{-6}$ | noncoding | 8.53 | 0.00 | - |
| rs6099115 | 20:56366084 | *FAM210B* | T | C | EUR+AFR | 0.34 | 4.54 | | $5.75 \times 10^{-6}$ | missense | 8.70 | 0.00 | likely_benign |
| rs765832505 | 13:113534120 | *TMCO3* | T | A | EUR | 0.02 | -1.16 | 0.27 | $8.22 \times 10^{-6}$ | missense | 5.68 | 0.04 | likely_benign |

### Gene-based analysis

In EUR, while there were no genome-wide significant findings, the gene-based analysis identified suggestive genes (*p*<1×10^-4^). These included *SLC22A10* (*p*=1.55×10^-5^; Supplementary Table 10) with the cumulative burden resulting from the category of rare synonymous variants (MAF≤0.001), *TMCO3* (*p*=2.79×10^-5^) with the cumulative variant burden from all LoF+missense variants (MAF≤0.5), and *FAM90A1* (*p*=6.06×10^-5^) evaluated by the aggregated effects of rare (MAF≤0.001) LoF+missense+synonymous variants. In AFR, we identified genes *DHX58* (*p*=5.20×10^-5^, LoF+missense, MAF≤0.001), *CHRND* (*p*=5.99×10^-5^, synonymous, MAF≤0.01), *GLDN* from the burden of LoF+missense+synoymous variants (*p*=6.77×10^-5^ for MAF≤0.01; *p*=7.00×10^-5^ for MAF≤0.5), *PLAT* (*p*=7.00×10^-5^, LoF+missense, MAF≤0.01), and *H1-4* (*p*=8.59×10^-5^, synonymous, MAF≤0.001) (Supplementary Table 11). In the cross-ancestry meta-analysis, *CHRND* was the most significant association (*p*=4.39×10^-6^, synonymous, MAF≤0.01; Supplementary Table 12), almost reaching the Bonferroni-corrected significance threshold. The other top genes include *QPCTL* (*p*=4.25×10^-5^), *COL3A1* (*p*=7.95×10^-5^), and *GPHB5* (*p*=9.50×10^-5^) (Supplementary Table 6).

**Table 3.** Top associated genes (*p*<1×10^-4^) in the gene-based tests. EUR, European ancestry; AFR, African ancestry.

| Gene | Category | MAF bin | Ancestry | P-value |
| --- | --- | --- | --- | --- |
| *CHRND* | synonymous | 0.01 | EUR+AFR | $4.39 \times 10^{-6}$ |
| *SLC22A10* | synonymous | 0.001 | EUR | $1.55 \times 10^{-5}$ |
| *TMCO3* | LoF+missense | 0.5 | EUR | $2.79 \times 10^{-5}$ |
| *QPCTL* | synonymous | 0.5 | EUR+AFR | $4.25 \times 10^{-5}$ |
| *DHX58* | LoF+missense | 0.001 | AFR | $5.20 \times 10^{-5}$ |
| *CHRND* | synonymous | 0.01 | AFR | $5.99 \times 10^{-5}$ |
| *FAM90A1* | LoF+missense+synonymous | 0.001 | EUR | $6.06 \times 10^{-5}$ |
| *GLDN* | LoF+missense+synonymous | 0.01 | AFR | $6.77 \times 10^{-5}$ |
| *GLDN* | LoF+missense+synonymous | 0.5 | AFR | $7.00 \times 10^{-5}$ |
| *PLAT* | LoF+missense | 0.5 | AFR | $7.00 \times 10^{-5}$ |
| *COL3A1* | LoF+missense | 0.5 | EUR+AFR | $7.95 \times 10^{-5}$ |
| *H1-4* | synonymous | 0.001 | AFR | $8.59 \times 10^{-5}$ |
| *GPHB5* | LoF+missense | 0.5 | EUR+AFR | $9.50 \times 10^{-5}$ |

GO and pathway enrichment analyses of the nominally significant genes (*p*<0.05) identified from the meta-analysis of gene-based tests identified the main clusters of metabolic regulation and tissue development. Clusters were defined based on the major biological themes that represent the similarities between GO terms/pathways. Among the largest cluster annotated as “metabolic regulation”, the enriched terms comprised those related to stimulus response and cell communication, among which the roles of opioid signals have been well characterized^50-52^ (Figure 3, Supplementary Table 13). Within-ancestry, gene-set enrichment analyses demonstrated nominally significant GO terms in alcohol metabolic process, and KEGG pathways related to vasopressin-regulated water reabsorption and hexose stimulus (Figure 3, Supplementary Tables 14-15).

**Figure 1.**
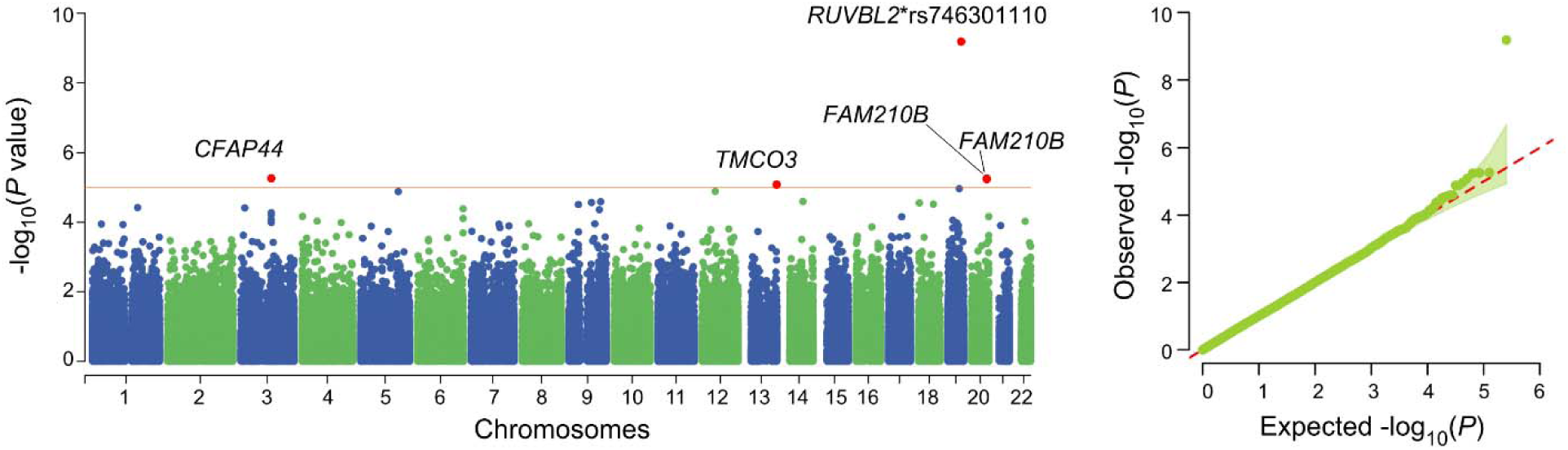
Cross-ancestry meta-analysis for single-variant association results. A total of 253,281 variants have been analyzed. The red horizontal line indicates a significance threshold of suggestive association (*p*=1×10^-5^). *N*_case_= 2,185, *N*_control_= 1,705.

**Figure 2.**
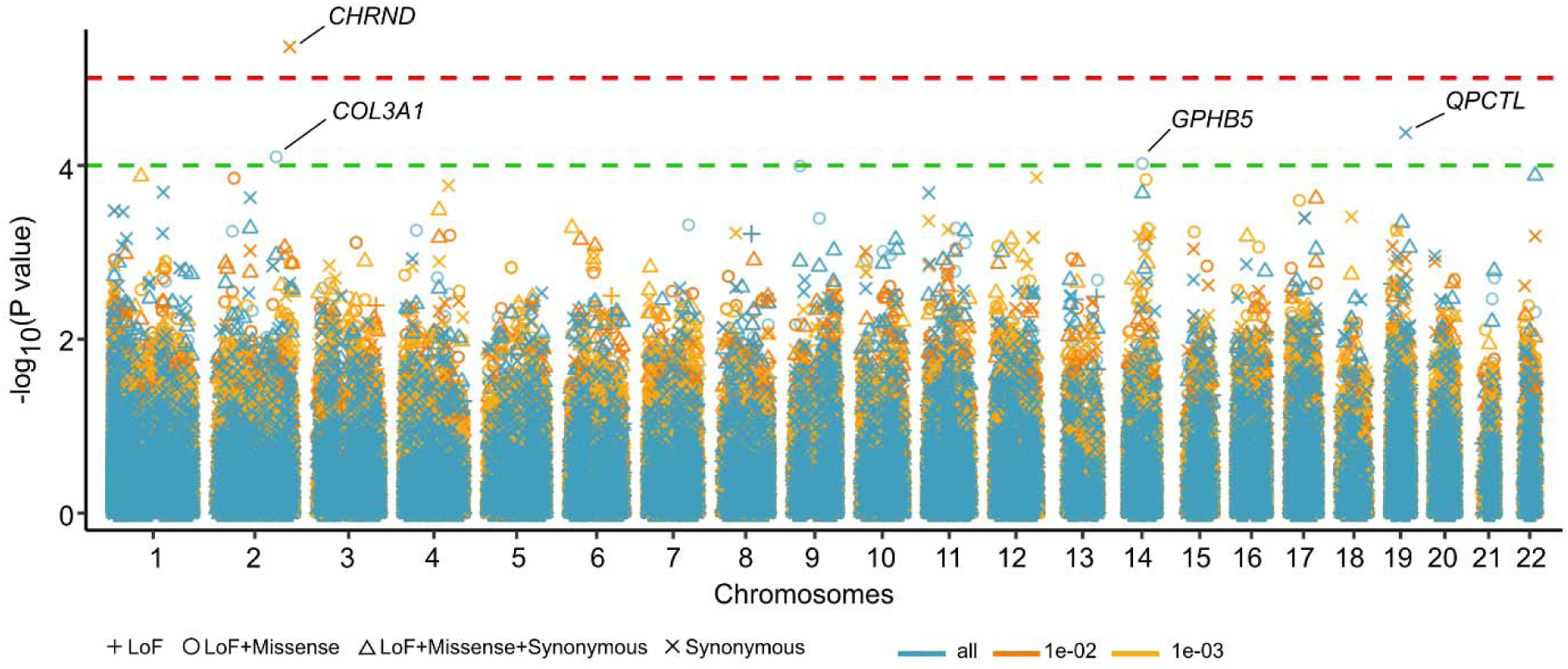
Meta-analyses of gene-based associations. Manhattan plot for meta-analyses of gene-based tests across the combinations of four variant categories and three MAF cutoffs. Burdens arising from different variant groups were represented by shape, with different MAF cutoffs separated by color. The red dashed line indicates the *p*-value threshold of 1×10^-5^, the green dashed line indicates a *p*-value of 1×10^-4^. Gene markers passing the threshold of 1×10^-4^ were marked.

**Figure 3.**
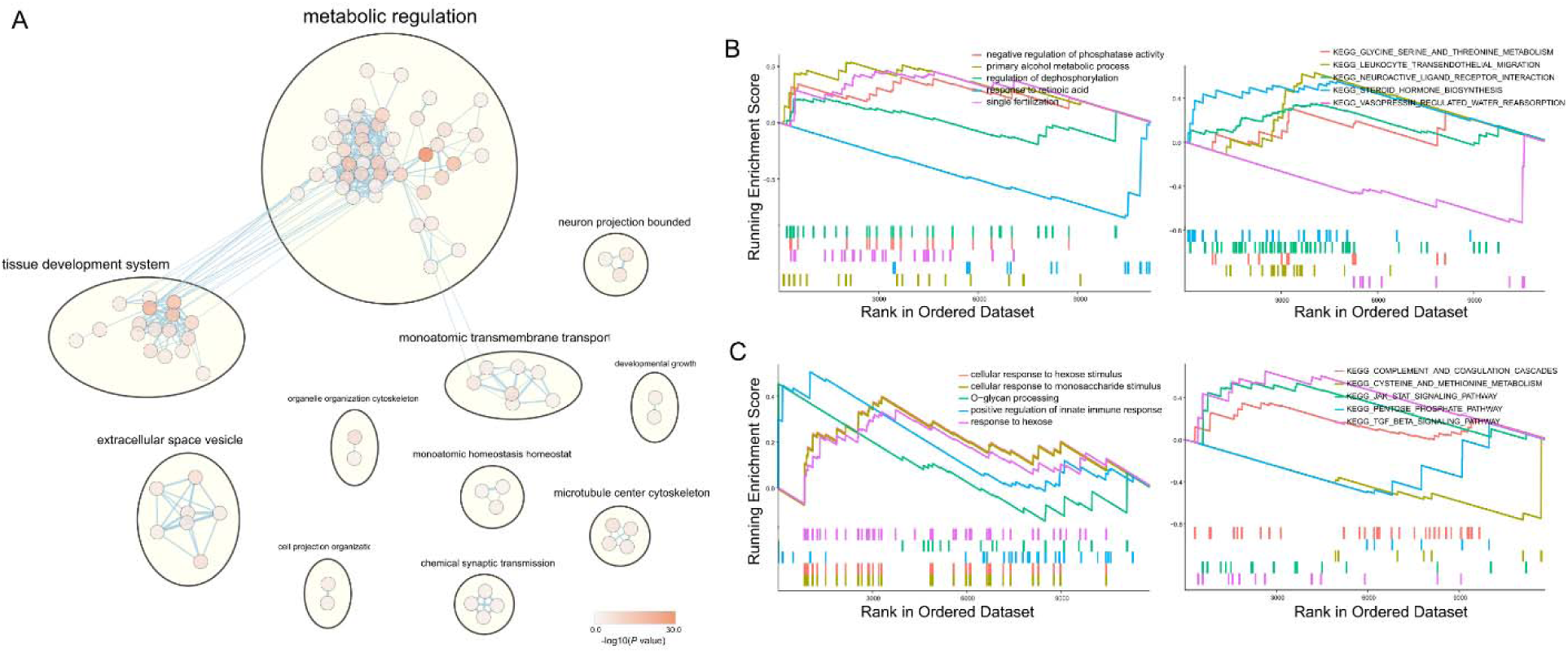
Gene set enrichment analyses for meta-analysis of gene-based results. **(A)** Integrative enrichment network of the genes derived from meta-gene-based analyses (*p*<0.05) across GO biological processes, molecular function, cellular component and Reactome database. **(B-C)** Top five enriched terms from gene-set enrichment analysis (GSEA) for selected genes (*p*<0.05) from gene-based associations across GO biological processes (left) and KEGG pathways (right) for EA **(B)** and AA **(C)**.

## Discussion

OD has enormous adverse health and economic consequences worldwide^4,5^. We identified a novel loss of function rare variant in the *RUVBL2* (RuvB Like AAA ATPase 2) gene that was genome-wide significant, and multiple suggestive variants. Gene-based collapsing tests identified gene associations attributable to rare variants and driven by the burden of loss-of-function and missense variants.

Understanding the molecular mechanisms of OD could help in prevention efforts and to treat OD by developing novel drugs or repurposing existing ones. OD is moderately heritable^9^. As with other complex psychiatric disorders, many genes contribute to the etiology of OD, each with a small effect size^19^. Before the application of GWAS in this area, the most studied candidate genes include opioid receptor genes^8^ (*OPRM1* encodes *µ*-opioid receptor, *OPRD1* encodes δ-opioid receptor), *DRD2* (dopamine receptor D2), and *BDNF* (brain-derived neurotrophic factor; reviewed in ref^18^). A functional coding variant rs1799971 (Asn40Asp) in *OPRM1* has been studied extensively to understand its function in relation to OD and was confirmed in a moderately powerful GWAS^13^. Since then, larger studies confirmed the association with the *OPRM1* locus repeatedly and discovered more than a dozen risk loci associated with OD^15-17^. In this study, *OPRM1*\*rs1799971 is nominally significant in EUR (*p*=1.72×10^-2^) but not in AFR (*p*=0.90), consistent with previous observations^13^ and reflecting the lower number of subjects in the present study compared to those where significant association was identified.

WES and whole-genome sequencing have increasingly been used in human genetic studies, as rare variants or variants not genotyped in GWAS can explain part of the genetic architecture of complex diseases. Only a few WES studies of substance use or use disorders have been conducted^55,56^, including ours^28^. However, whereas there are no prior published WES studies of OD, and this multi-ancestry study comprising 3,890 participants from the Yale-Penn cohort is the first.

We identified an exome-wide significant (and also genomewide-signficant) rare variant in the *RUVBL2* gene (rs746301110, *p*=6.59×10^-10^) in EUR participants. This variant is a low-frequency INDEL (ATAGA/-, MAF<0.01) in EUR and is common (MAF >0.01) in most non-European populations (data from gnomAD v4.1.0^57^). Rs746301110 has potential functional consequences evidenced by its CADD score of 27, which predicts a high likelihood of deleteriousness. *In silico* prediction using SpliceAI predicts a splice acceptor variant with a probability of 1 (Table 2). The *RUVBL2* (RuvB Like AAA ATPase 2) gene encodes a DNA helicase essential for homologous recombination and DNA double-strand break repair, thus it plays an important role in transcriptional regulation^58^ and cancers^59,60^. GWAS have identified associations between variants in *RUVBL2* and biometric traits including multiple blood protein levels^61^. Further research is warranted to ascertain the biological mechanism that may connect genetic variation at this gene locus with OD.

Other findings with suggestive evidence (*p* <1×10^-5^) were identified in genes *NEIL2*, *CFAP44*, *FAM210B*, and *TMCO3*. *NEIL2* (Nei like DNA glycosylase 2) encodes a DNA glycosylase and also plays a role in DNA break repair. Variants within this gene were associated with biometric traits^62^ and lipid level interactions with alcohol consumption^63^. Two variants in high-LD in the *FAM210B* gene which encodes a mitochondrial membrane protein were identified in the cross-ancestry meta-analysis, one is in the intron region and the other is a missense coding variant. AlphaMissense predicts the missense variant to be likely benign. Another missense in the *TMCO3* (transmembrane and coiled-coil domains 3) gene was identified, with a predicted pathogenicity of likely benign. Interestingly, variants in these genes were associated with several biometric and anthropometric traits, recorded in the GWAS Catalog^64^.

The most significantly associated gene from the gene-based tests is *CHRND* (Cholinergic Receptor Nicotinic Delta Subunit) (*p*=4.39×10^-6^) in the cross-ancestry analysis, which was mainly driven by the results in AFR (*p*=5.99×10^-6^). This gene encodes the delta subunit of the nicotinic acetylcholine receptor, which mediates synaptic transmission at the neuromuscular junction. Defects in this gene are a cause of multiple muscle-related disorders^65^. A burden of synonymous rare variants (MAF <0.001) in *SLC22A10* was observed in European samples (*p*=1.55×10^-5^). Variants in *SCL22A10* have been reported to be associated with blood and urine biomarkers in the UK Biobank^62^. Convergent evidence was obtained in European samples from both the single-variant and gene-based associations in the *TMCO3* gene, with the burden of Lof+missense variants (*p*=2.79×10^-5^).

This study has limitations, and paramount among these is small sample size compared to that required for well-powered complex trait genomic discovery. Increasing the study sample by incorporating biobank-level data has proven to be a useful way to identify novel risk variants associated with SUDs^15,17,66^. However, there is limited sequence data from individuals with OD in publicly available biobanks. Thus, this study of <4,000 subjects lacks ideal power but it is the first study of its kind. Another limitation is that WES only covers the coding regions of the genome, and we therefore cannot uncover the part of the genetic architecture of OD that exists in non-coding regions. Larger studies are required to identify more associations and to confirm the ones we present here.

Our study includes roughly equal numbers of EUR and AFR subjects, avoiding a bias that is common in genetics studies. Focusing on the coding regions is essential to understanding the variant effect on proteins, as these are among the best targets for drug development, and we successfully identified significant risk variants and genes.

## Data Availability

All data produced in the present study are available upon reasonable request to the authors.

## Acknowledgments

We want to acknowledge the participants in the Yale-Penn cohorts. The authors are supported by grants from the National Institutes of Health (NIH) (R01 DA12890, R01 AA026364, R01 AA11330, R01 DA037974, P30 DA046345, P50 AA012870, U54 AA027989, RM1 HG011558, and R01 AA030056) and the U.S. Department of Veterans Affairs (1I01CX001849 and I01 BX004820).

